# Multisystem inflammatory syndrome in children related to COVID-19: A systematic review

**DOI:** 10.1101/2020.08.17.20173641

**Authors:** Hoste Levi, Van Paemel Ruben, Haerynck Filomeen

## Abstract

**Importance:** In April 2020, multiple reports of an association between a hyperinflammatory, Kawasaki-like condition and SARS-CoV-2 were published and termed as pediatric inflammatory multisystem syndrome (PIMS) or multisystem inflammatory syndrome (MIS). A thorough characterization of this syndrome (demographics, presentation, diagnosis, and outcome) is currently lacking.

**Objective:** We aimed to perform a systematic review of published cases of this novel multisystem inflammatory syndrome in children associated with COVID-19.

**Evidence review:** A literature search of Pubmed, Embase, BioRxiv, MedRxiv and COVID-19 specific research repositories (Cochrane COVID-19 Study Register and the World Health Organization (WHO) COVID-19 Global Research Database) was conducted from December 30^th^, 2019 to June 30^th^, 2020. Publications describing inflammatory syndromes associated with COVID-19 were included. Of 333 unique publications, 229 records were excluded based on title and abstract. After screening the full text, 40 observational studies and case reports were included, comprising 687 cases (published between May 9^th^, 2020 and June 30^th^, 2020).

**Findings:** In contrast to classic Kawasaki disease, epidemiological enrichment for adolescents (median age 9 [6.0-12.3]) and ethnic minorities (35.8% black and 24.5% Hispanic/Latino) was observed. There was a male predominance (59.1%). Apart from obesity (24.4%), pre-existing conditions were infrequent. The majority suffered from gastrointestinal (87.2%) and cardiocirculatory (79.2%) manifestations. Respiratory symptoms (51.2%) were less frequent. Over half of patients (56.3%) presented with hemodynamic shock, and critical care interventions were often necessary (inotropics (56.5%), mechanical ventilation (22.9%), non-invasive ventilation (30.6%), extracorporal membrane oxygenation (ECMO;4.5%)). Anti-SARS-CoV-2 IgG and RT-PCR were positive in respectively 69.4% and 36.7%. Eleven deaths were reported (1.6%). The RCPCH case definition proved to be most comprehensive comprising all single cases. In contrast, WHO and CDC MIS definitions are more stringent, with the CDC case definition often missing severe cases requiring intensive care (n = 33 out of 95 cases).

**Conclusions and Relevance:** This novel pediatric multisystem hyperinflammatory condition, associated with COVID-19, is characterized by a severe and heterogeneous disease spectrum. Despite frequent intensive care interventions, mortality rate was low and short-term outcome favorable. Long-term follow-up of possible chronic complications and additional clinical research, to elucidate the underlying immunological pathogenesis and possible genetic predisposition is crucial.

**Key points:** *Question:* How is the novel pediatric multisystem inflammatory condition associated with coronavirus disease 2019 (COVID-19) characterized?

*Findings:* This systematic review of 40 studies, comprising 687 cases, represents the heterogeneous spectrum of this novel pediatric disease related to COVID-19, including contrasting features with previously-described hyperinflammatory conditions. Adolescents and particular racial/ethnic minorities are affected more. Gastrointestinal and cardiocirculatory manifestations are often found, along with critical care interventions. Nevertheless, only 11 deaths are reported.

*Meaning:* This novel condition has variable severity but good short-term outcome. Uniform case definitions are required to guide future (preferably controlled) research on epidemiological clustering, immunopathology, and long-term prognosis.

## Introduction

Severe acute respiratory syndrome coronavirus 2 (SARS-CoV-2), causing coronavirus disease 2019 (COVID-19), led to a pandemic health crisis within a few months’ time^1–3^. Severe COVID-19 and associated mortality has been highest in elderly persons and adult patients with comorbidities, such as cardiovascular disease, diabetes mellitus, and chronic lung disease^4–6^. Since the outbreak, COVID-19 was generally described as asymptomatic or mild in children, causing few pediatric hospitalizations and minimal mortality^7–10^.

Since April 2020, several countries from Europe and North America reported on young patients with a severe multisystem inflammatory syndrome associated temporally with SARS-CoV-2. The initial descriptions exposed important clinical heterogeneity, partially overlapping with features of Kawasaki disease (KD) or toxic shock syndrome (TSS), but nevertheless clearly distinct from such known inflammatory conditions^11,12^. In contrast with acute COVID-19, a significant proportion of children were reported with severe or fatal disease^11,13–17^. Since its description, this disease entity is most often referred to as pediatric inflammatory multisystem syndrome temporally associated with SARS-CoV-2 infection (PIMS-TS) in Europe^18,19^ or multisystem inflammatory syndrome in children (MIS(-C)) in the USA^20,21^.

At present, it is pivotal to optimize the characterization and the diagnostic criteria of this pediatric multisystem inflammatory syndrome related to COVID-19. To date, the scattered reporting of cases provides insufficient insight in the full clinical spectrum, epidemiological and immunological features, therapeutics and prognosis. Hence, we performed a systematic review, to describe the diagnostic criteria and the clinical manifestations of this novel COVID-19-associated phenotype in children.

## Methods

Original studies describing cases meeting the definition of PIMS-TS or MIS(-C) by the Royal College of Paediatrics and Child Health (RCPCH)^19^, World Health Organization (WHO)^21^ or Centers for Disease Control and Prevention (CDC)^20^, were eligible for inclusion (eMethods 1). Primary outcome analysis focused on epidemiological, clinical, and prognostic parameters.

A search strategy was designed with keywords combining the pediatric population, COVID-19 and hyperinflammatory presentations (eMethods 2), including articles published from December 31th, 2019 to June 30th, 2020. Electronic databases were searched (PubMed, Embase), including pre-print (BioRxiv, MedRxiv) and COVID-19-specific research repositories (Cochrane COVID-19 Study Register and WHO COVID-19 Global Research Database). The reference lists of included studies were considered as an additional source.

After duplicate removal, two reviewers (LH/RVP) independently applied the inclusion and exclusion criteria, first, by screening titles and abstracts and, second, by examining full texts. LH extracted data using a standardized form, while RVP cross-checked for correctness and completeness. Any disagreement was resolved by FH. The Preferred Reporting Items for Systematic Reviews and Meta-Analyses (PRISMA) checklist guided study selection and extraction. Risk for bias on observational studies^22^ and levels of evidence^23^ were assessed (LH) with verification (RVP). Prior to conducting the review, the protocol was published (PROSPERO CRD42020189248). Data was analyzed with R v3.6.3 (eMethods 3).

Cohort studies and studies reporting single case data were analyzed separately. Proportions were calculated on the sum of cases of studies reporting on the variable, except for rare conditions (e.g. death), which was calculated on the total group. Unfavorable outcome was defined as presence of coronary dilatation/aneurysm, shock, death, need for mechanical ventilation, extracorporal membrane oxygenation (ECMO), renal replacement therapy, inotropes or PICU admission.

## Results

### Study characteristics

The search strategy yielded 474 records. After removing duplicates, 333 unique publications were screened on title and abstract of which 229 were excluded. One hundred and four full-text articles were assessed for eligibility. Forty studies were included for the systematic review (Figure 1). In general, risk of bias was low (eTable 1), despite short follow-up.

**Figure 1:**
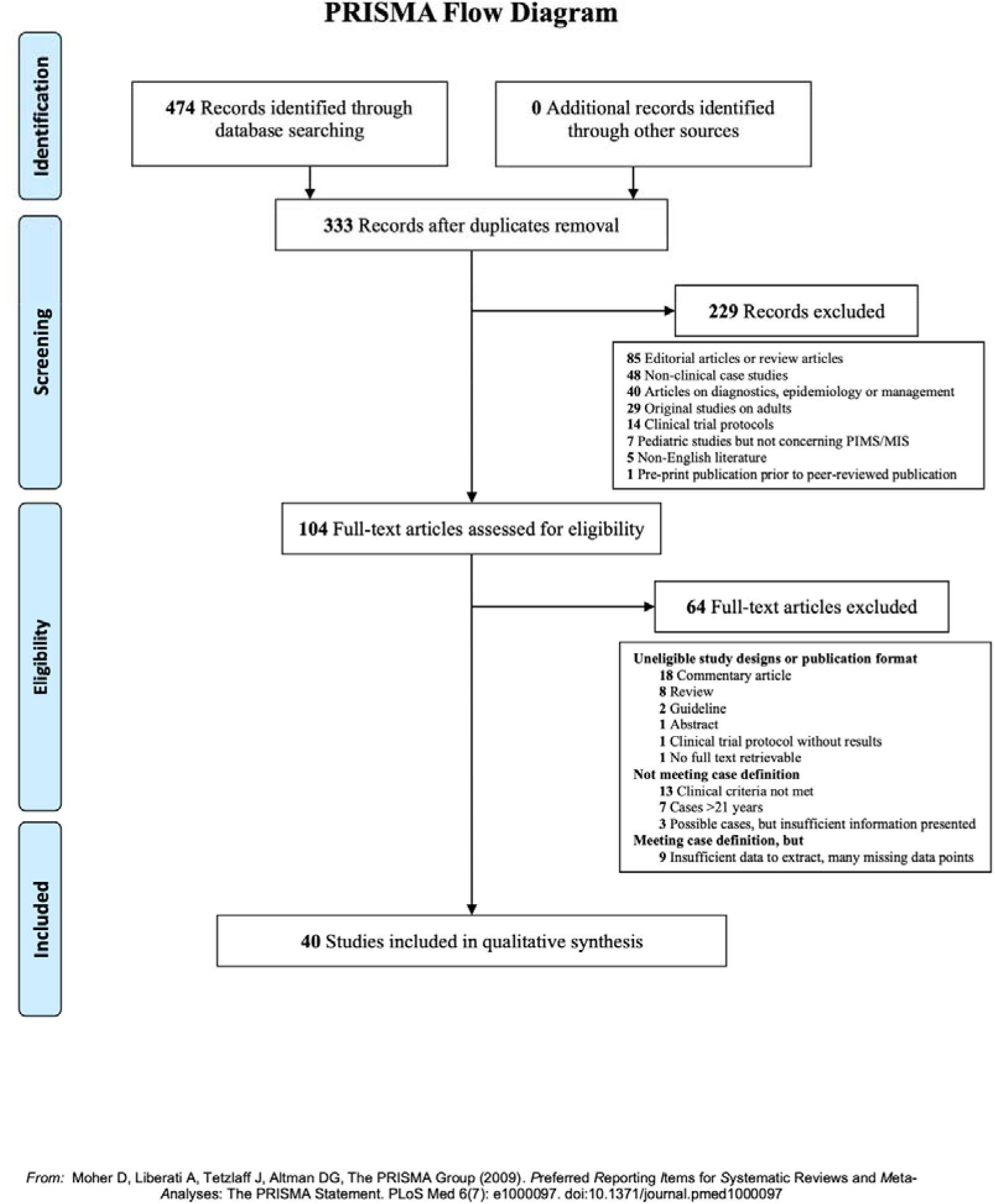
Preferred Reporting Items for Systematic Reviews and Meta-Analyses (PRISMA) flow diagram

All studies were published after May 9^th^ 2020 and presented observational data from single case reports^24-39^ or case series (2-186 cases per publication)^11–17,40-55^ (Figure 2-Table 1). All studies were non-controlled, although two publications^15,43^ used historical cohorts as a reference population. Studies were mostly conducted in the United States (n=18)^13,14,38,42,45–47,53,55,56,17,24,26,29,31–33,36^, Franc (n=6)^40,41,43,48,52,54^ or the UK(n=5)^11,15,16,44,50^. Reports from non-Western countries were rare^25,34,35,37,39^.

**Table 1:**
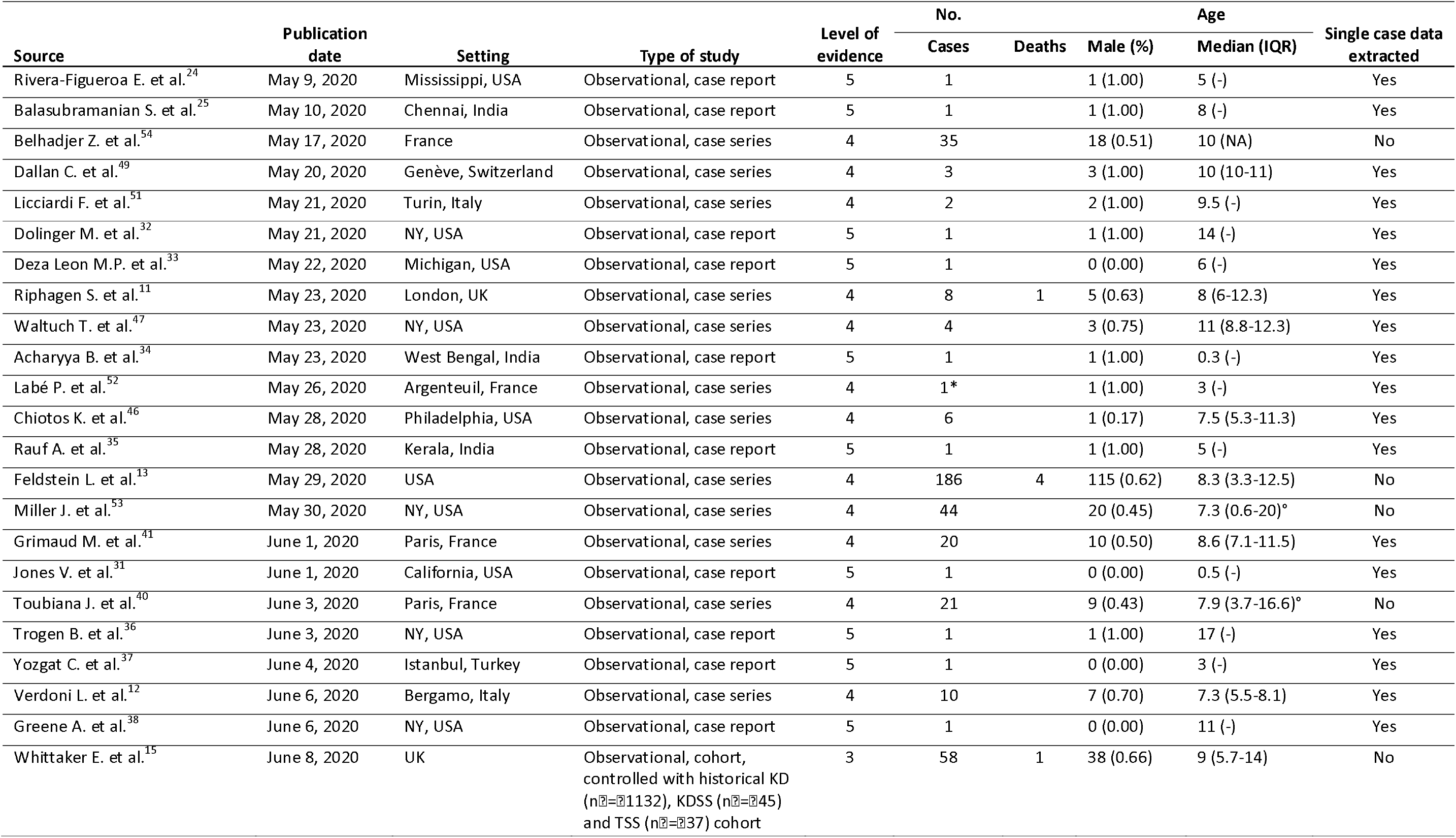
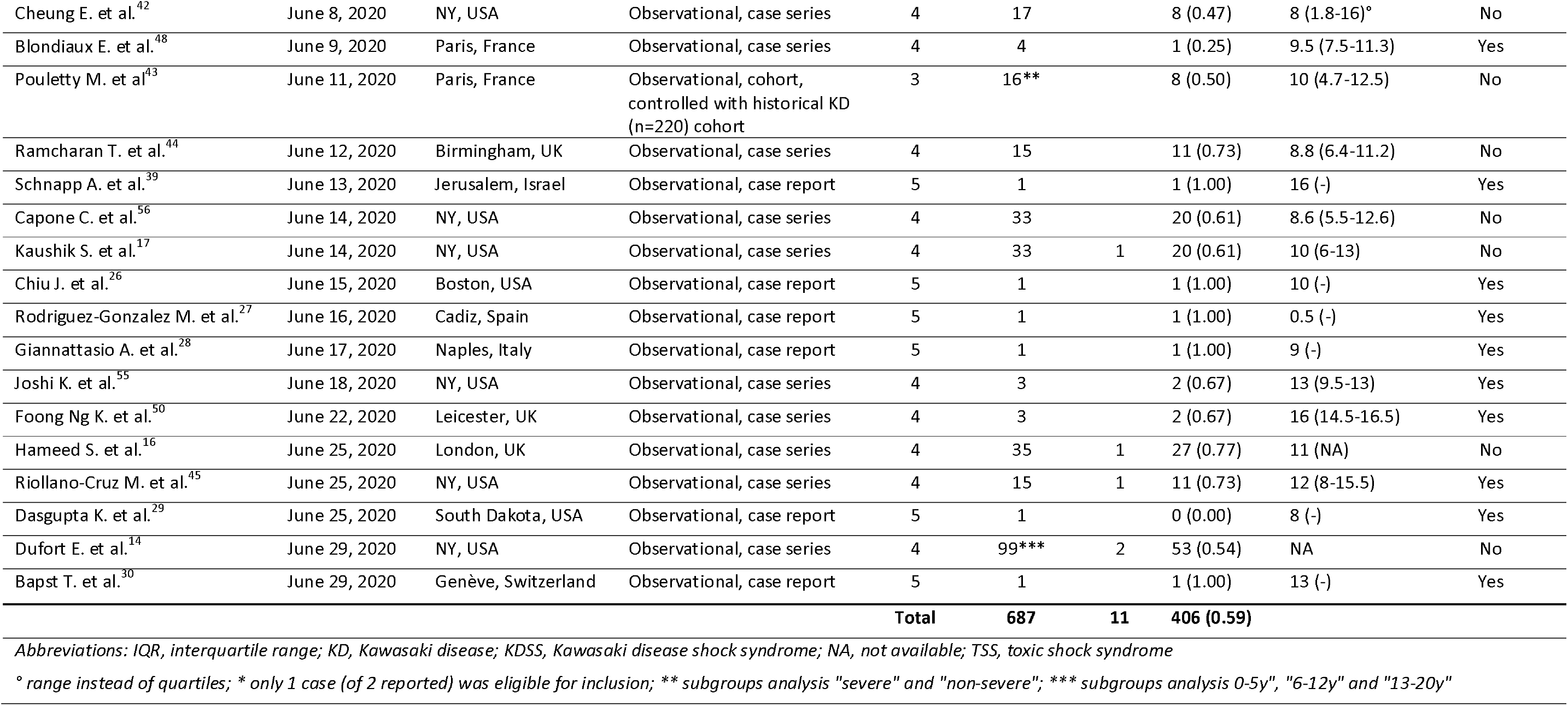
included studies in the systematic review

**Figure 2:**
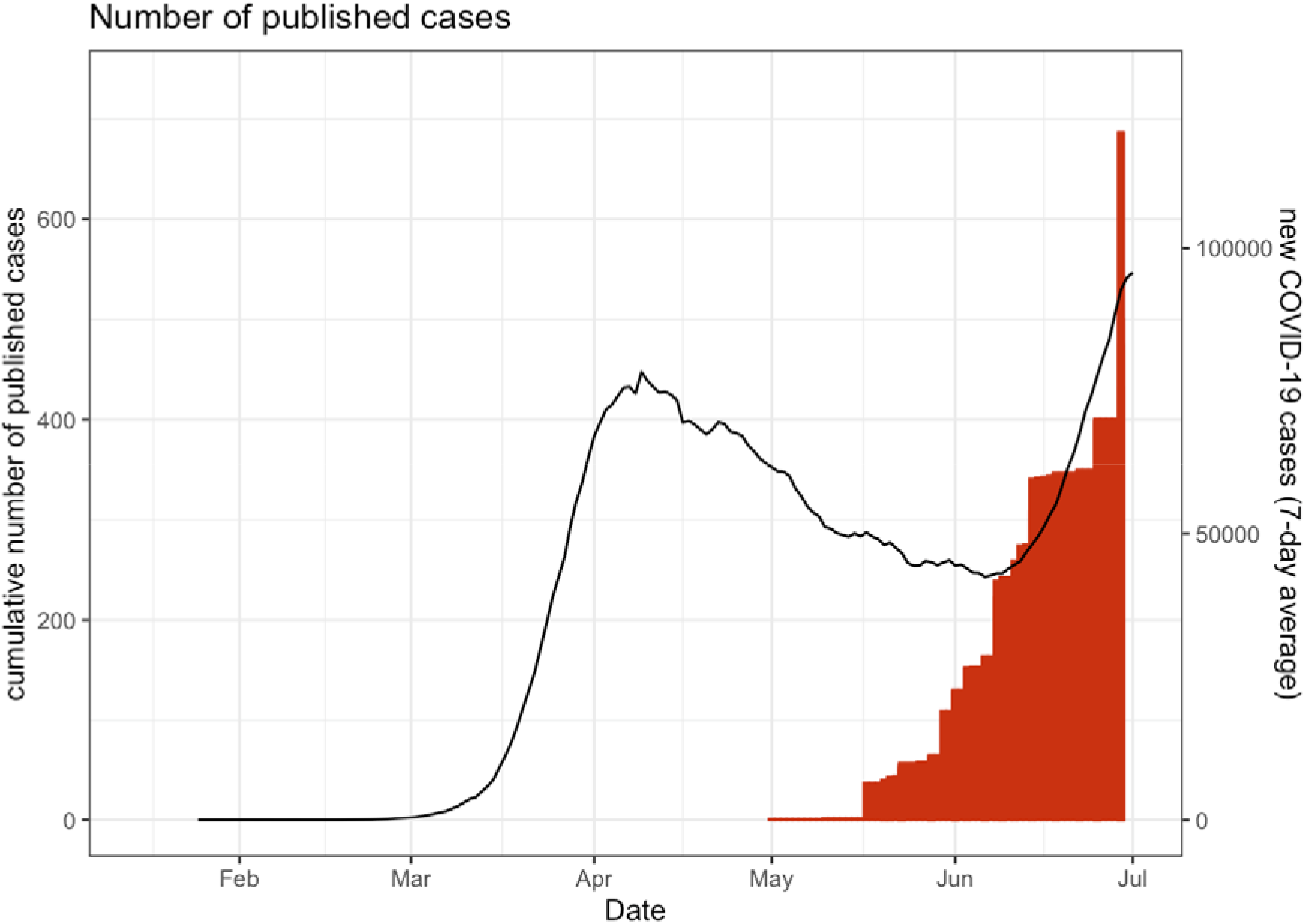
Cumulative number of published PIMS-TS/MIS(-C) cases (bars) in relation to total reported COVID-19 cases, according to data from Johns Hopkins Coronavirus Resource Center (lines; 7-day rolling average). The four countries (France, Italy, UK and US) with most included cases were plotted.

### Demographics

In total, 687 patients with PIMS-TS/MIS(-C) were reported with detailed patient information (single case data) available for 95 patients (14.0%)^11,12,32-39,41,45,24,46–52,55,25–31^. Seventeen patients (2.5%) were reported in duplicate, although the corresponding manuscripts^13,16,17^ did not provide sufficient information to filter for unique data.

Among single cases, a median age of 9 (IQR 6.0-12.3) was found (Figure 3A). A median age of at least 8 was present in 9/12 cohorts (419/493 cohort patients)^13,14,16,17,42–44,54,56^, Remarkably this median age was significantly higher compared to non-COVID-19 KD cases (median 2.0-2.7 years)^15,43^. Additionally, a clear male predominance (406/687;59.1%; Figure 3B) was found.

**Figure 3:**
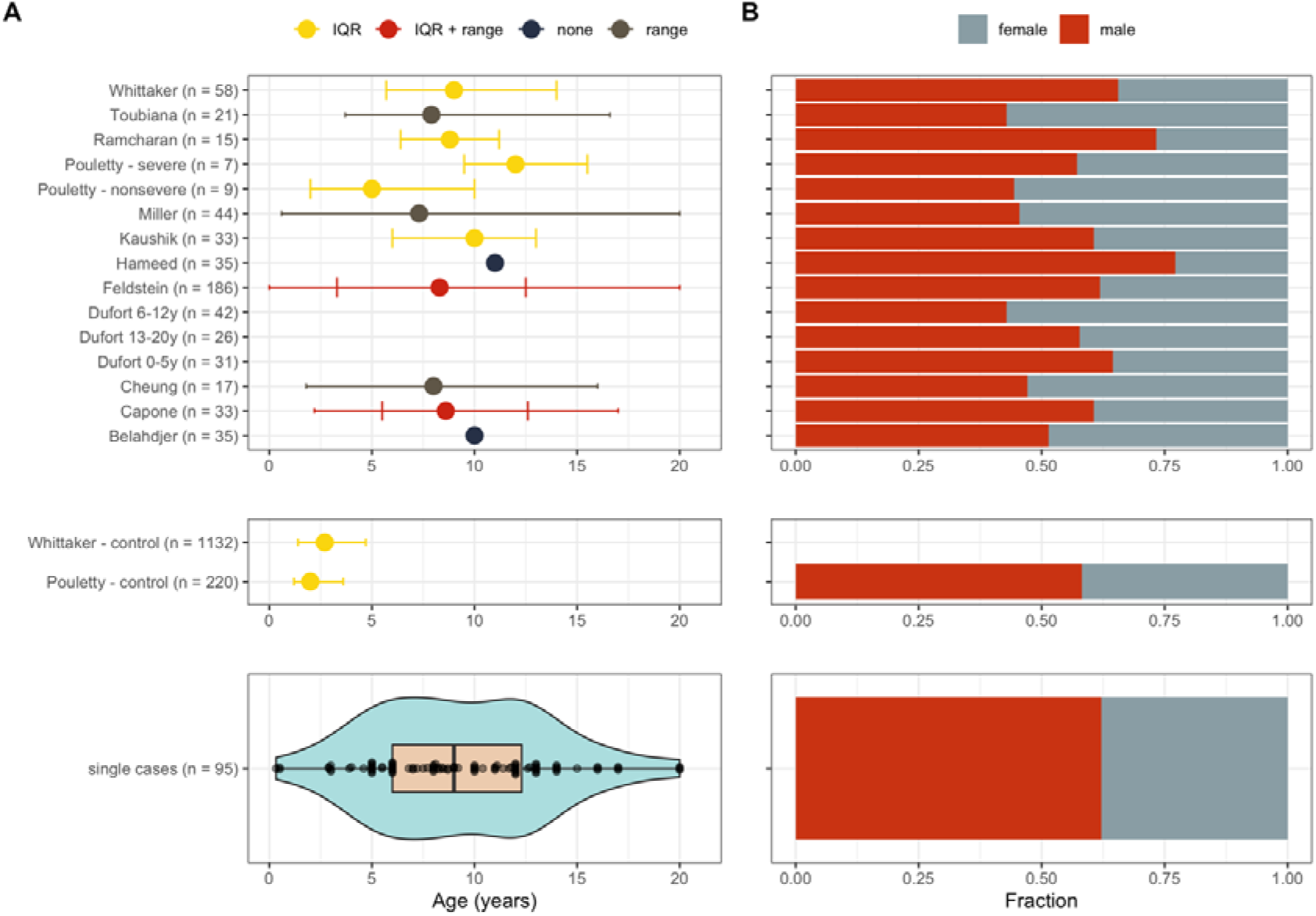
Demographic characteristics of all included studies. (A) Age distribution, (B) Fraction of males and females in each study. IQR = interquartile range.

PIMS-TS/MIS(-C) cases were frequent black (176/491; 35.8%), followed by patients of white race (132/491; 26.9%) and mixed/other/unknown (126/491; 25.7%)^11,13,45,46,49,50,53,56,14,15,17,24,29,40,42,44^. Hispanic/Latino was reported in 77/314 (24.5%)^11,13,24,29,45,46,49,50,56^. Co-morbidities were infrequent, and mainly consisted of respiratory diseases, including asthma (29/687;4.2%)^15,17,56,36,42,43,45,47,49,54,55^ or chronic lung disease (14/687;2.0%)^14^. Overweight (BMI>25 kg/m^2^ or >85th percentile for age/sex) was found in 132/541 (24.4%)^11,13,53-56,14,17,36,43,45,46,49,50^. Immunodeficiency (10/687;2.7%)^13^ and cardiovascular disease (8/687;1.2%)^13,17,55^ were less frequent.

### Clinical presentation

Fever was documented in nearly all patients (683/687;99.4%)^11,12,27–36,13,37,46,14,47–55,15–17,24–26^; often during at least 5 days (239/687;34.8%). The majority (520/596;87.2%)^11,12,30–33,35–40,13,41–50,14,51–56,16,24,26–29^ presented gastro-intestinal symptoms, mostly abdominal pain (210/329;63.8%)^11,14,36,38,39,41,45–50,15,51,53,55,17,24,28–30,32,35^, vomiting (204/350;58.3%)^11,14,47-51,53,15,17,29,36,40,41,45,46^, and djarrhea (175/350;50.0%)^11,12,36,40,45–48,50,51,53,55,14,15,17,24,26,27,29,35^. Half of cases (248/484;51.2%)^12,13,31–34,36,37,42,43,45,47,15,48–57,24–30^ showed respiratory symptoms, including upper respiratory tract symptoms (44/252; 17.5%)^14,15,54,55,27,29,31,33,45,47,50,52^, dyspnea (52/227;22.9%)^14,17,49–51,54,55,24,25,27,31,33,45,46,48^ and (multiple) radiological infiltrates (74/195;37.9%)^12,16,42,44–47,49,50,52,55,17,25,27,29–31,33,36^. Cardiovascular manifestations were found in 79.4% of patients (258/325)^11,12,31–39,41,13,45–53,55,24–30^. Tachycardia (176/210;83.8%)^11,14,37,38,41,43,45,47–51,25,55,26,27,29,31,32,35,36^, hemodynamic shock or hypotension (270/480;56.3%)^11,12,29,32,33,35,36,38,41–44,14,45,47–51,53–56,15,57,16,17,24–27^, myocarditis (123/266;46.2%)^14,16,41,43,48,51,26,27,29,30,33,35,36,40^and mild or moderate decreased left ventricular ejection fraction (LVEF between 30 and 55%;169/414;40.8%)^11,12,44–46,48,50,51,54–56,13,17,33,35,36,38,39,42^ were frequently observed cardiovascular abnormalities. Severe complications such as LVEF less than 30% (32/435;7.4%)^11,12,56,13,17,40,42,44,45,48,54^ coronary dilatation (z-score between 2.0-2.5;62/511;12.1%)^11,13,46,47,50,54,56,15,16,34,40,42–45^ or aneurysms (z-score above 2.5;29/373;7.8%)^12,14,15,40,42,47,49,50,54,56^ were found in a minority of cases. Pericardial effusion was frequently found (100/420;23.8%)^12,13,47,48,50,51,54,17,24,26,40,42–45^. Acute kidney injury was described in 90/535(16.8%)^11,13,49,53,55,56,14,15,35,36,38,39,41,46^ Only 9 cases revealed thrombotic complications^11,13,16,17^,including 2 splenic infarctions^16^ and 3 ischemic strokes secondary to cerebral hemorrhage during EC MO^11,16,17^.

The presence of KD criteria (eMethods l)^58^ was assessed among included cases. Polymorphous exanthema (364/639;57.0%)^11,12,29–34,37–40,13,41–43,45–51,14,52–54,15–17,24–26^ and non-puruent conjunctivitis (344/639;53.8%)^11,12,29–31,34,35,37,40–43,13,45–54,14–17,24–26^ occurred most. Only 15 of 95 single cases fulfilled criteria for classic KD (15.8%)^12,24,52,25,29,31,34,37,47,48,51^. A quarter (24/95;25.3%) fulfilled 2 or 3 of the KD criteria in combination with prolonged fever^11,12,47,48,26,30,33,35,38,41,45,46^ Similarly, incomplete KD was reported in 59/239 (24.7%) of cohort cases^13,40,42,44^. Although shock was frequently reported in single cases (80/95;84.2%)^11,12,36,38,41,45–51,24,55,25–27,29,32,33,35^; shock with classic KD was rare (8/95;8.4%)^12,24,25,29,34,47,48,51^.

### Assessment and diagnosis

Increased inflammatory markers were often documented (Figure 4), including C-reactive protein (median 256 mg/l [IQR 183-335] in single cases)^11,12,32–39,41,45,24,46–50,52,55,25–31^; ferritin (939 μg/l [525-1478.5])^11,12,36–38,45–47,49–51,55,24–28,32,33,35^ and interleukin-6 (235 pg/ml [IQR 121.0-374])^27,28,32,38,43,45,47,50^. Notable, patients exhibited substantially higher inflammation compared to historical KD cohorts.

**Figure 4:**
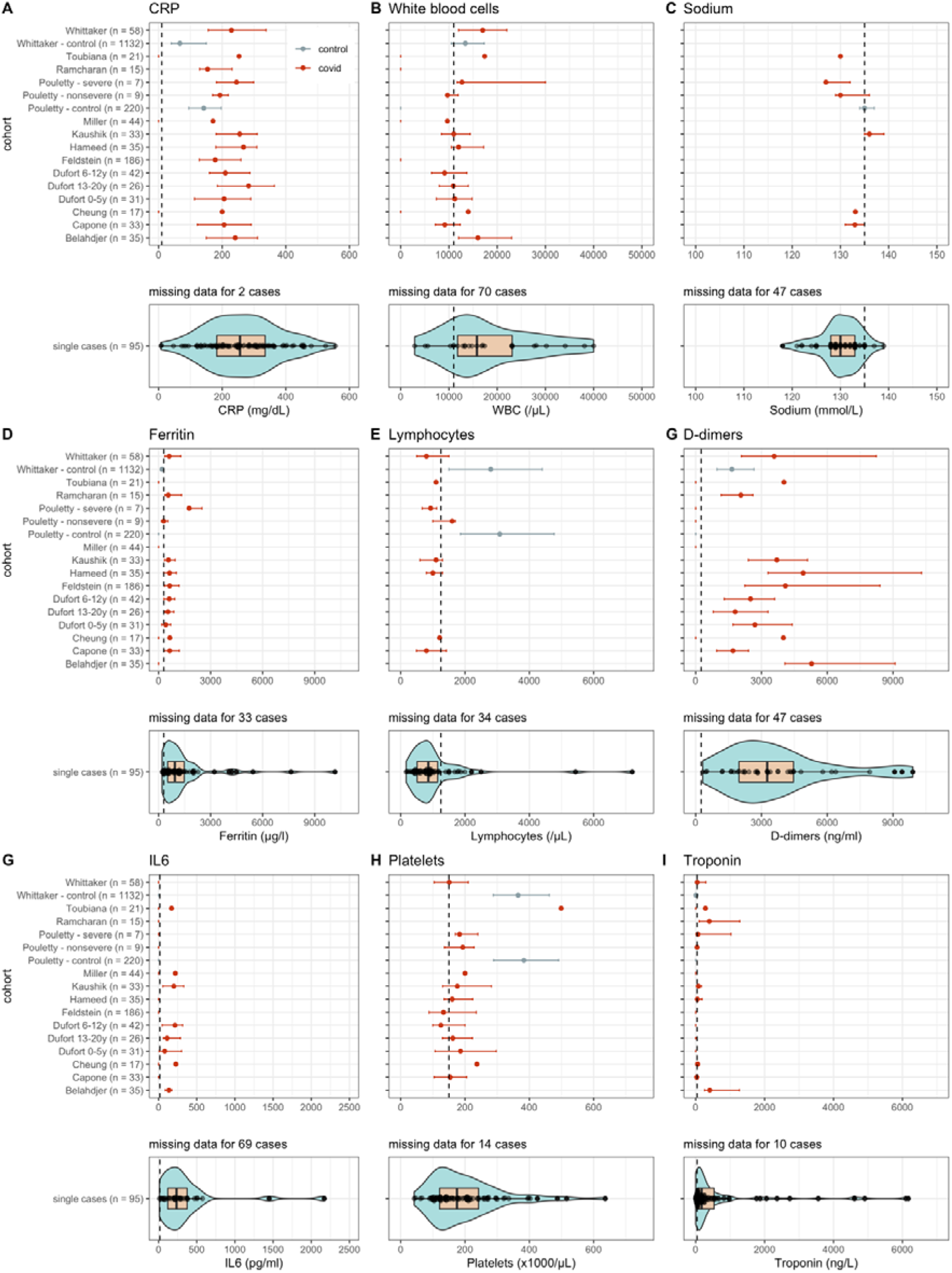
Laboratory tests values and distribution for each study. Error bars correspond to the interquartile range. Dashed vertical line equals the upper limit of normal (CRP, white blood cells, ferritin, D-dimers, IL6 and troponin) or the lower limit of normal (sodium, lymphocytes and platelets). For studies that report multiple values for the same test, the maximum (CRP, white blood cells, ferritin, D-dimers, IL6 and troponin) or the minimum (sodium, lymphocytes, platelets) was used. “Covid” (red line) equals values corresponding to the COVID-19 related hyperinflammatory syndrome; “control” (gray line) equal values corresponding to the Kawasaki control populations as described by Pouletty and Whittaker.

Although white blood cell count was frequently increased, (WBC 15800/μl [IQR 11800-23110])^24,25,38,47,49,50,52,55,26–29,33–35,37^, lymphocytopenia was common (860/μl [IQR 510-1150]^12,26,39,41,46–51,27–30,33,35–37^, contrasting with historical KD cohorts^15,43^ (median lymphocytes 2800-3080/μl). Thrombocytosis is a typical KD sign and serves as a laboratory criterium for incomplete KD^59^. However, most PIMS-TS/MIS(-C) presented reduced to normal thrombocytes (platelets below 15000/μl in 34/81;42.0%, while platelets above 450,000/μl in only 5/81;6.2%)^11,12,34–37,41,45–47,49,50,24–30,33^.

Besides inflammatory parameters, coagulation markers were substantially upregulated, including increased D-dimer and fibrinogen^11,26,41,45–47,49,50,55,27–29,32,36–39^. Furthermore, markers of myocardial injury such as troponins and brain natriuretic peptide (BNP) were often elevated^11,12,36–38,41,45–50,24,55,26–30,33,35^ Hyponatremia (130 mmol/l [IQR 128-133])^12,24,46,48,25,26,29,31,33,35,36,41^ was frequent, in contrast with historical KD patients (135 mmol/l [134-137])^43^.

### SARS-CoV-2 testing

Current or recent SARS-CoV-2 infection was assessed with RT-PCR (nasopharyngeal or fecal swab) and/or serological assays (IgG/lgM/lgA). The majority of the patients were IgG positive (272/392;69.4%)^12,14,43,46-51,54,56,15,16,27,28,30,39-41^. IgM (5.7-65.7%)^16,54^ and IgA positivity (25/35;71.4%)^54^ were documented in only two and one cohort, respectively. All single patients being IgA (18/95; 18.9%)^30,41,49^ or IgM positive (7/95;7.4%)^12,28,48,51^, had detectable IgG antibodies as well. Only 252/687 (36.7%) had positive respiratory RT-PCR^12,13,32–34,36,38,40–44,14,45,46,49,50,53–56,15–17,24–26,31^ Positive fecal RT-PCR was rare (6/225;2.7%)^16,41,43,53,54^. Close contacts with COVID-19 were registered in 127/418 (30.4%)^11,12,44,49,51,52,54,13,17,28,30,34,40,42,43^. Thirteen out of 95 cases were negative for both PCR and serology^11,12,29,35,37,41,52^, of which 9 additionally had no known COVID-19 contact^11,12,29,35,37,41^.

As an inclusion criterium, all cases in this review corresponded to at least one of the recognized case definitions^19–21^. The RCPCH definition, not requiring proven or probable SARS-CoV-2 infection, was most comprehensive, as it comprised all single cases. Although more stringent concerning clinical manifestations and the relationship with SARS-CoV-2, the WHO definition nevertheless included 98% (9 not assessable), missing only 2 mild cases. In contrast, the CDC definition comprised only 67% (7 not assessable) of single cases and neglected on 24 cases with unfavorable outcome (see Methods). All single cases needing ECMO (n=3)^11,33,45^ or who died (n=2)^11,45^ corresponded to all three definitions.

### Therapeutic management

Three quarter of patients (500/652;76.7%) received intravenous immunoglobulins (IVIG)^11,12,33–35,37,38,40–44,13,45–48,50–55,14,56,15,17,24,25,29,31^, and multiple IVIG doses were needed in 67/500 (13.4%)^13,40,43,44,46,50,54,56^. Systemic corticosteroids were prescribed in 353/625 (54.1%)^11,12,38–46,48,13,50,51,53-56,14,15,17,27-29,35^. Acetylsalicylic acid was reported in 110/197 (55.8%) of which 26/110 (23.6%) received high, anti-inflammatory dosages (SO-lOOmg/kg/day)^11,12,42–46,48,50,56,24,29,31,33–35,37,40^. Heparin (268/407;65.8%) was a frequent anti-thrombotic^11,13,45,47,54–56,17,27,28,32,33,36,42,44^.

One hundred and fourteen cases (114/687; 16.6%) were treated with biopharmaceuticals, including anakinra (52/687;7.6%)^13,15,56,17,43,45–47,49,53,54^, interleukin-6 inhibitors (tocilizumab/siltuximab; 51/687;7.4%)^13,17,56,25,27,38,42,43,45,47,55^, and to a lesser extent, infliximab (11/687; 1.6%)^11,15,32,56^. Remdesivir (11/687;1.6%) was rarely prescribed^17,38,45,55^.

Among patients receiving critical care, inotropics were given to 363/643 (56.5%)^11,12,33,35,38–45,13,46–51,54,–56,14-17,26,27,29^ Mechanical and non-invasive ventilation was initiated in 157/687 (22.9%)^11,13,41,43–50,53,14,54–56,15–17,27,33,39,40^ and 37/234 (30.6%)^11,14,45-1,54,55,17,24,25,33,35,41,43,44^ respectively A relative high rate of ECMO (31/687;4.5%)^11,13-17,33,45,54^ was reported. Renal replacement therapy (9/687;1.3%) was less frequently provided^11,13,49,53^.

### Prognosis and outcomes

Intensive care admission was common (456/596 (76.5%) with median duration of 4 days [IQR 3.75-8] in single cases and 4-7 days in cohorts^11,13,43-46,49,50,54-56,14,16,17,27,35,40-42^. The median time of total hospitalization was 8 days (IQR 7-12) in single cases and 4-12 days in cohorts^13,17,50,54,56,24,35,36,42,44–46,49^. Residual cardiac dysfunction, often reported as residual decreased left ventricular ejection fraction at discharge or follow-up, was present in 20/244 (8.2%)^17,42–44,54,56^. No other residual morbidity was reported.

Eleven deaths were described (11/687;1.6%)^11,13-17,45^. One patient was less than 1 year old^14^, 4 were aged 5-12 years^13,14,17,45^ and 4 others were older than 13 years^11,13^. The majority was male (5/7) and black (5/6)^11,13,17^. All reported deaths presented with shock and/or myocardial dysfunction, needing inotropics and/or mechanical circulatory support^11,13-17,45^. ECMO was initiated in 8/11 of fatal cases, of which 3 died of hemorrhagic cerebral infarction^11,16,17^. Co-morbidities among fatal cases were obesity (4/11;36.4%)^11,13^, asthma (n=1)^13^ and multiple neurological conditions (n=1)^13^.

## Discussion

Overall, children and adolescents with COVID-19 exhibit mild or asymptomatic disease. Only limited reports of complicated or fatal COVID-19 in children are published^7,10,60,61^. Although some immunological hypotheses are presented^62-65^, hitherto, obvious elucidation on why children display a milder COVID-19 phenotype is lacking.

At the end of April 2020, while over 3 million individual SARS-CoV-2 infections were reported, a relative sudden emerge of clusters of children presenting a life-threatening hyperinflammatory disorder with multisystem involvement, legitimately, prompted an international alert. Noteworthy, during the previous 2 months, more than 650 individual cases with PIMS-TS/MIS(-C) have been reported in peer-reviewed literature, and, subsequently, systematically reviewed herein. Currently, 6 months after the onset of the pandemic, several countries are still struggling with widespread SARS-CoV-2, requiring continuous and evidence-based updates on the COVID-19 spectrum, in particular concerning complicated disease courses. In this context, we performed a systematic review on PIMS-TS/MIS(-C) to appropriately characterize its clinical presentation, treatment and prognosis.

The findings in this review confirm the heterogeneous spectrum in PIMS-TS/MIS(-C). The majority of PIMS-TS/MIS(-C) patients presented gastrointestinal symptoms, even without (prior) respiratory symptoms, despite SARS-CoV-2 displaying respiratory tract tropism^66^. Cardiovascular manifestations, including severe circulatory failure and myocardial involvement requiring intensive care, burdens PIMS-TS/MIS(-C) substantially, and was dominantly present in all deceased patients. Nevertheless, the majority of patients (98.4%) survived the acute phase of PI MS-TS/M IS(-C). Noteworthy, male predominance and overrepresentation of ethnic minorities (black, Hispanic or Latino), as well as the absence of reports from Asian countries, is observed in this review. Apart from obesity, co-morbidities are strikingly missing, also among fatal cases. So far, underlying factors such as genetic predisposition, prior infections or immunizations contributing to PIMS-TS/MIS(-C) vulnerability are unclear.

Comparing with historical KD cohorts^15,43^, patients with PIMS-TS/MIS(-C) are substantially older (typically adolescent), and represent more systemic inflammation, lymphocytopenia, thrombocytopenia, myocardial injury and coagulopathy. Of the PIMS-TS/MIS(-C) cases fulfilling classic KD criteria, half presented with shock, in contrast with non-COVID-19 associated KD shock syndrome with an incidence rate of only 3.3-7% of KD cases^67,68^. Moreover, coronary dilatation (12.1%) and aneurysm formation (7.8%) are more prevalent than in appropriately treated KD (~5%), as well as mortality rates, typically less than 0.1% in KD (versus 1.6 % in PIMS-TS/MIS(-C))^58^. Despite some overlapping features, this review confirms that PIMS-TS/MIS(-C) is a distinct entity from KD, KD shock syndrome, toxic shock syndrome, or immune dysregulation syndromes.

Severe COVID-19 might be related to host immune overdrive and unbounded cytokine release^69,70^. In contrast with severe adult COVID-19, notable respiratory symptoms are lacking in PIMS-TS/MIS(-C), and primary respiratory failure does not seem a dominant cause for ICU admission. The clinical presentation of PIMS-TS/MIS(-C) with evidence for systemic vasculitis, multisystem involvement, and hypercoagulative state however resembles the pathophysiological mechanism as macrophage activation syndromes, but remains insufficiently studied on a molecular level. Besides some observational data on cytokines, no thorough immunological characterization of PIMS-TS/MIS(-C) cases were found during the study period of this review. As such, further research efforts are required. Understanding of the involved immunological pathways might contribute to accurate therapeutic approaches that interfere with these dysregulated immune responses. Although abnormal coagulation parameters were frequently reported (D-dimers and fibrinogen as signs of disseminated intravascular coagulation), thrombotic or embolic events were rare, in contrast with adult severe COVID-19^71^.

Based on a seroconversion rate of more than two thirds, this review confirms the highly probable association with recent SARS-CoV-2 infection in PIMS-TS/MIS(-C). In addition, a delay of PIMS-TS/MIS(-C) cluster with 4-6 weeks after the peak of COVID-19 in the community has been documented previously, suggestive of antibody-driven pathogenesis. The seropositivity rate moreover contrasts with concurrent seroprevalence studies, showing limited seroconversion in the pediatric community^72^. Although the presence of unbounded, active SARS-CoV-2 infection has not been thoroughly assessed, active viral replication in the pathogenesis of PIMS-TS/MIS(-C) seems unlikely with positive RT-PCRs in only 1/3 cases.

The true incidence of PIMS-TS/MIS(-C) remains unknown and, in the current crisis, a notification bias might be present. In absence of comprehensive surveillance studies among the pediatric population, the proportion of SARS-CoV-2 infected children subsequently suffering from PIMS-TS/MIS(-C) can only be estimated. A better understanding of affected age groups and associated risk factors is thus necessary.

Ultimately, this review has several limitations. In particular, we were unable to collect individual data of all patients and the single case data are obtained from case reports and case series. We did not contact the authors of included studies for additional information or insights in their data as we believed this would significantly delay the reporting of this pressing data. Due to the nature of included studies, these reports are moreover enriched for severe disease course (e.g. shock being more frequent in single cases than cohorts). Furthermore, only two^15,43^ of the included studies contain a historical control population. In addition, the initial association of COVID-19 could have triggered a reporting bias. It is reasonable that this might result in overdiagnosis of PIMS-TS/MIS(-C), which can be appreciated from the single case proportions meeting the different case definitions. The sensitivity and specificity of recognized case definitions should prospectively be assessed. This review currently favors the use of the WHO MIS definition, being specific (e.g. concerning the association with COVID-19), while remaining 98% sensitive. In future studies, we furthermore recommend including a control population with similar disease characteristics in whom a recent or close contact with COVID-19 is excluded, in order to further characterize PIMS-TS/MIS(-C).

As this review was conducted while PIMS-TS/MIS(-C) has only been described since a few months, inevitably, delayed complications or long-term effects were not yet assessed. Despite high proportions of patients requiring intensive care, overall prognosis remained good, with the caveat of undervaluing long-term prognosis to date.

Because the relatively small number in the single case cohort and many lacking data in larger cohorts, formal statistical testing was not conducted. As such, the findings of this review should be interpreted as descriptive and exploratory. Due to the retrospective nature of included studies, and not all studies reporting all variables, we were unable to collect sufficient data to develop a clinical prediction model for disease course or treatment response. To date, there is a lack of randomized controlled trials concerning PIMS-TS/MIS(-C). As a surrogate, systematic reviewing of observational data might contribute to the expertise required and rapidly identify gaps in knowledge. Updating the data set of this review, based on future publications in the coming months, might consecutively provide answers to these needs.

## Conclusions

A novel hyperinflammatory condition with severe multisystem involvement has been described in children and adolescents in the midst of the COVID-19 pandemic (PIMS-TS/MIS(-C)). This review systematically assessed its epidemiological enrichment for adolescents and racial and ethnic minorities, its probable association with SARS-CoV-2, its clinical heterogeneous presentation with frequent gastrointestinal manifestations and circulatory failure including myocardial injury, and lastly, its overall good prognosis with absence of short-term complications despite frequent need for critical care interventions. Further epidemiological, clinical, immunological and genetic research is urgently needed, as well as long-term follow-up studies of PIMS-TS/MIS(-C) patients.

## Data Availability

All raw data will be made available on Github (https://github.com/rmvpaeme/PIMS_MISC_SR) after peer review.
The processed data has been added as a supplementary file.

## Author contributions

All authors contributed to the study conception and design. The review process and author contributions are outlined in Methods. The first draft of the manuscript was written by Levi Hoste and all authors commented on previous versions of the manuscript. All authors read and approved the final manuscript.

## Acknowledgements

L. H. was funded by the VIB Grand Challenge project.

R.V.P was funded by a predoctoral fellowship from the Research Foundation Flanders (FWO).

